# Predicting the cumulative medical load of COVID-19 outbreaks after the peak in daily fatalities

**DOI:** 10.1101/2020.09.03.20183384

**Authors:** Claudius Gros, Roser Valenti, Lukas Schneider, Benedikt Gutsche, Dimitrije Marković

## Abstract

The distinct ways the COVID-19 pandemics has been unfolding in different countries and regions suggest that local societal and governmental structures play an important role not only for the the baseline infection rate, but also for the short-term and long-term reaction to the outbreak. We propose to investigate the question of how societies as a whole, and governments in particular, modulate the dynamics of a novel epidemic using a generalization of the SIR model, the controlled SIR model. We posit that containment measures are equivalent to a feedback between the status of the outbreak and the reproduction factor. Short-term control of an outbreak, corresponds in this framework to the reaction of governments and individuals to daily cases and fatalities. The reaction to the cumulative number of cases or deaths, and not to daily numbers, is captured in contrast by long-term control. We present the exact phase space solution of the controlled SIR model and use it to quantify containment policies for a large number of countries in terms of short- and long-term control parameters. We find increased contributions of long-term control for countries and regions in which the outbreak was suppressed substantially together with a strong correlation between the strength of societal and governmental policies and the time needed to contain COVID-19 outbreaks. Furthermore, for numerous countries and regions we identified a predictive relation between the number of fatalities within a fixed period before and after the peak daily fatality count which predicts the cumulative medical load of COVID-19 outbreaks that should be expected after the peak. These results suggest applicability of the proposed model not only for understanding the outbreak dynamics, but also for predicting future cases and fatalities once the effectiveness of outbreak suppression policies is established with sufficient certainty.

**Author summary:** The country specific dynamics of the COVID-19 pandemics has been suggests that local societal response and governmental structures are critical both for the baseline infection rate and the short-term and long-term reaction to the outbreak. Here we investigate how societies as a whole, and governments, in particular, modulate the dynamics of a novel epidemic using the controlled SIR model, a generalisation of a standard compartmental model used for modelling the dynamics of infectious diseases. We posit that containment measures correspond to feedback between the status of the outbreak (the daily or the cumulative number of cases and fatalities) and the reproduction factor.

We present the exact phase space solution of the controlled SIR model and use it to quantify containment policies for a large number of countries in terms of model parameters corresponding to long- and short-term control. Furthermore, we identified for numerous countries a relationship between the number of fatalities within a fixed period before and after the peak in daily fatalities. As the number of fatalities corresponds to the number of hospitalised patients, the relationship can be used to predict the cumulative medical load, once the effectiveness of outbreak suppression policies is established with sufficient certainty.

## 1 Introduction

Epidemic outbreaks differ widely with respect to the parameters defining their dynamics and societal impact, such as infection rate, fatality rate, and the rate of critical cases requiring hospitalization. In order to gauge the effectiveness and suitability of policies enacted for outbreak containment one needs therefore to discern the outbreak parameters. However, in novel outbreak the precise assessment of any of these quantities and a prediction of disease spreading is in practice a notoriously difficult task [1]. Outbreak case data at early stages is often both noisy and biased [2], which implies that core epidemiological parameters cannot be estimated with sufficient precision. This is a hard limitation of any epidemics-forecasting approach, as small differences in disease dynamical properties can lead to drastically different outcomes [3].

However, in spite of these limitations, it is safe to assume that both individuals and governments will react to a spread of a new infectious disease. Given the severity of the COVID-19 outbreak [4], it is unsurprising that the raising case and fatality numbers not only forced governments to impose lock-down measures [5,6], but also motivated people to avoid travelling and mass gatherings [7]. Hence, to understand the dynamics of COVID-19 outbreak, we propose to model the feedback of spontaneous societal and imposed governmental restrictions using a standard epidemic model that is modified in one key point: the reproduction rate of the virus is not constant, but evolves over time alongside with the disease in a way that leads to a ‘flattening of the curve’ [8]. The basis of the proposed model is the SIR (Susceptible, Infected, Recovered) model, which describes the evolution of a contagious disease for which immunity persists substantially longer than the outbreak itself [9]. We extend the model by introducing a negative feedback loop between the severity of the outbreak and the initial reproduction rate *g*_0_. Our model contains two parameters, *α_X_* and *α_I_*, which control respectively the amount of long- and short-term epidemic control. The first, *α*_X_, quantifies the contribution of the cumulative case count X to the negative feedback loop. For the second parameter, *a*/, short-term control, the growth rate is reduced when the current number of active cases, I, is large. The resulting process is denoted the controlled SIR model.

The controlled SIR model draws its motivation from previous epidemiology modelling. One of the first evidences showing that human behaviour affects spreading dynamics [10,11], came from the study of measles epidemics [12]. Generalizations of the SIR model account for various effects of societal response to an outbreak, such as self-isolation [13], contact-frequency reduction and quarantine [14], changes in human mobility [15], together with the effects of geographic and societal networks [16], and of explicit feedback loops [17]. For a detailed analysis, epidemiology models can be extended to cover a range of additional aspects [18], with an example being the distinction between symptomatic and asymptomatic cases [19]. These kind of complex models are in general not accessible to an explicit analytic handling. It has also been questioned, whether detailed modelling leads to improved predictions [1,2], given that field data is inherently noisy.

The controlled SIR model was recently introduced in [20], where the authors analysed long- or short-term control separately, but not arbitrary mixtures. In contrast, we derive here an analytic solution for the controlled SIR model in the presence of both long-term and short-term control. Additionally, we show that the descriptions of COVID-19 field data is substantially improved when both short-term and long-term control are included. For data analysis and model validation, we use publicly available COVID-19 case and fatality counts for a wide range of countries and regions.

We find that the cumulative number of fatalities within a given period of a few weeks before the peak number of daily fatalities increases by 30% for the same period after the peak. Strikingly, this result is found to hold universally across all considered countries and regions. In contrast to the universality with regard to the increase in fatalities, substantial differences in the country-specific intrinsic reproduction factor and short and long-term control parameters are found. A comprehensive theoretical description based on an analytic solution of the controlled SIR model is given, together with a detailed validation (based on simulated data) of the statistical inference used for estimation of the country specific parameters. We also evaluate search-engine based measures quantifying the effectiveness of lock-down measures and the impact of structural factors (e.g. population density) on the infection rate and doubling time. Finally, we conclude that the controlled SIR model allows precise quantification of the outbreak dynamics, and provides a predictive framework for assessing the effectiveness of containment measures and future medical load.

## 2 Results

In the following we will use the symbol *X* to denote cumulative case counts, both for field data and for theory results. The number of new cases, which are typically reported in official COVID-19 datasets on a daily basis, we will denote with Δ*X*. We will add a time tag subscript d to denote reported counts on a specific day. In that case, the following sum rule holds: *X_d_* = ∑*_d″≤d_* Δ*X_d″_*. In analogy, we denote with F and *ΔF* cumulative and daily fatalities, respectively. Importantly, the model presented here is explicitly defined for one isolated epidemic outbreak. Our analysis focuses thus exclusively on the initial outbreak, as defined in more details in Data smoothing / peak definition.

### 2.1 Controlled SIR model

The logic of an infectious disease is described by the SIR model,

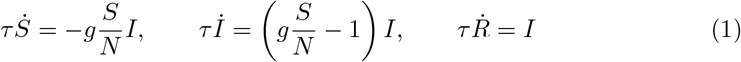

which takes the number of susceptible *S* = *S*(*t*), infected *I = I*(*t*) and recovered (removed) individuals *R* = *R*(*t*), as constituents variables [21]. The sum, *S* +1 + *R* = *N*, is assumed to be constant at all times *t*, as the population size *N* is approximately unchanged over the course of the the outbreak. In its basic formulation, the SIR model is characterized by a timescale, *τ*, and a dimensionless reproduction factor, *g*.

Note that the Eq. (1) describes an uncontrolled isolated outbreak in an environment that does not react to the disease. In reality, counter measures will be taken either spontaneously by the general public, or will be imposed by governmental institutions. As a result the reproduction factor will fall below its intrinsic value, which we denote with *g*_0_. We posit that reactions to the unfolding of the epidemic are based either on the current situation (the current active cases, *I*) or on the overall history of the outbreak (the total cases *X* = *N* − *S*). Formally, we can express this dependence as

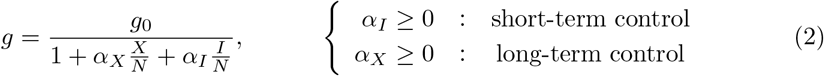

Note that the functional form in Eq. (2) parallels the law of diminishing return [22], which reflects the intuition that containment becomes progressively harder.

The controlled SIR model, Eq. (1) together with Eq. (2), can be solved exactly (see Sect. Exact solution of the controlled SIR model for more details). The exact solution corresponds to the following relation

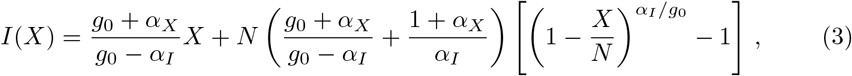

which we will use throughout the paper to illustrate the time independent phase-space representation of the data, as shown in Fig. 1. Note that both long- and short-term control reduce the severity of an outbreak with respect to the uncontrolled case, *a_X_* = *a_I_* = 0, however with distinct shapes of the phase space trajectories. The phase-space (XI) representation tends to be stretched for short-term control and parabola-like for long-term control [20].

**Fig 1.**
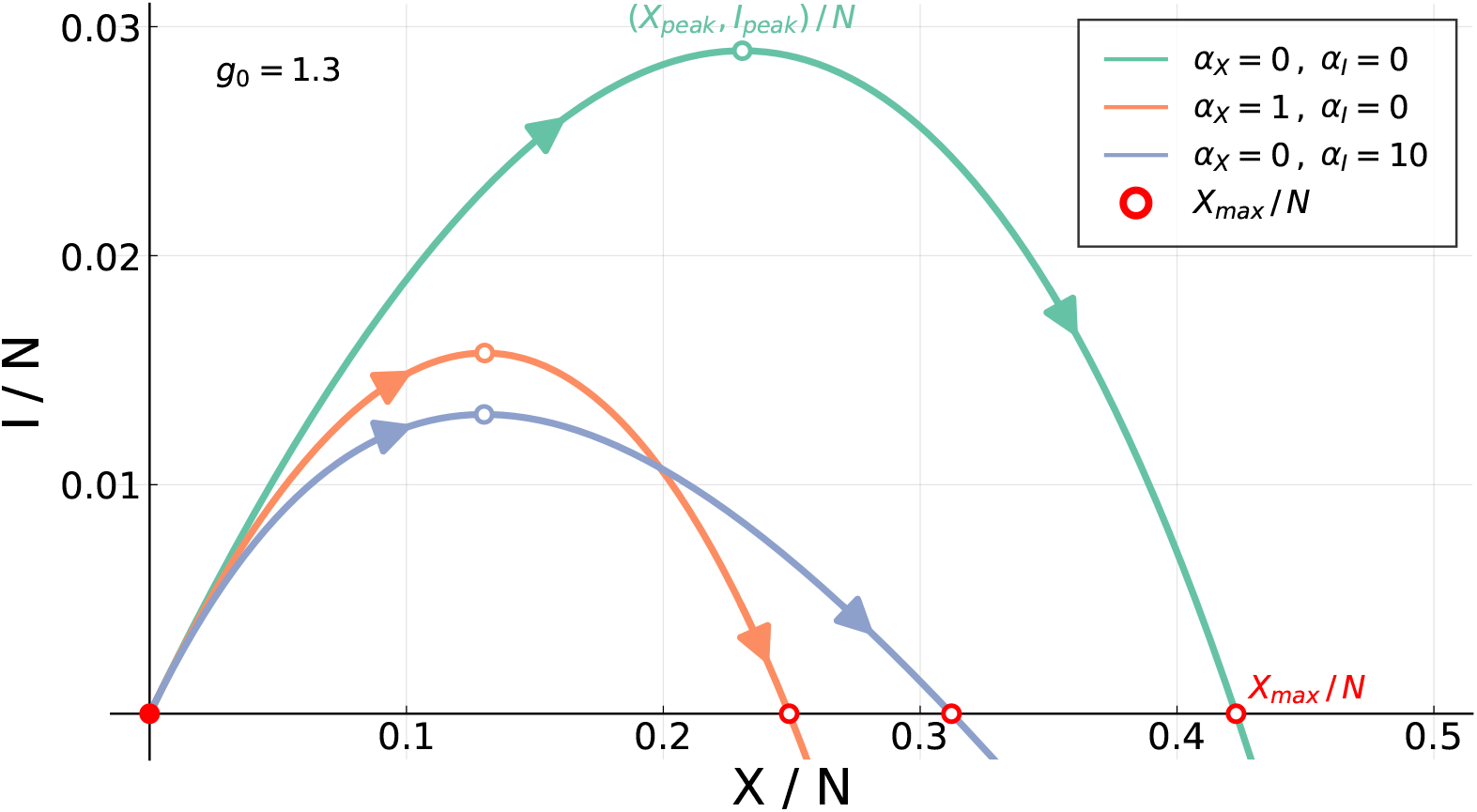
Short and Long-term control. Outbreaks are contained when either short-term or long-term control is present, see Eqs. (1) and (2). Long-term control (*α_X_* > 0, *α_I_* = 0) produces more symmetrically confined outbreaks than short-term control (*α_X_* = 0, *α_I_ >* 0).

The maximum of *I*, the peak rate *I*_peak_, is obtained for

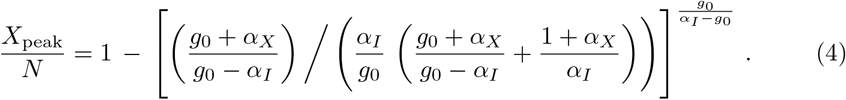

From (4) one obtains *I_peak_* via (3).

### 2.2 Data validation

As an example of the COVID-19 data examined we present in Fig. 2 the timeline of the outbreak for the US state. Also shown is a comparison of several publicly available data sources (see Sect. Data sources for details). For most daily values the John Hopkins and ECDC data agree, as illustrated in Fig. 2b for the case of Spain, Turkey and Germany. For the latter, the case counts published by the Robert Koch Institute have been added. Spain is a special case, as the official counting criteria did see a major revision end of May 2020 [23].

**Fig 2.**
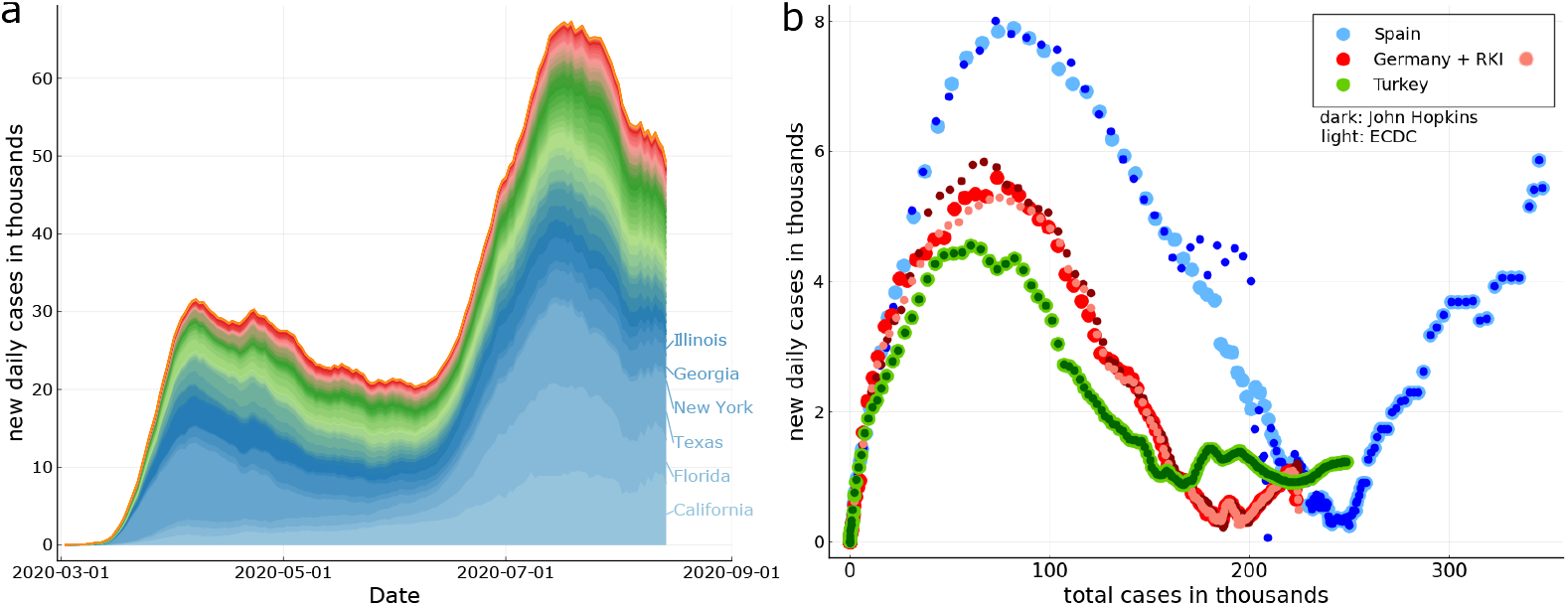
COVID-19 outbreak examples. *Left:* The timeline of the daily new infections for the US states. The curves are in part strongly asymmetric with respect to the time it take for the outbreak to build up and to recede. A second, prominent peak is present. *Right:* For the XI-representation daily counts are plotted as a function of total counts. For Spain, Germany and Turkey a comparison of ECDC (European Center of Disease Control), the John Hopkins, and the RKI (Robert Koch Institute, Germany) data. Seven day moving averages have been used. Note the substantial scattering of the later-stage COVID-19 data for Spain, which is due to changes of official counting protocols. For the data sources see Sect. Data sources.

A key focus of the present study concerns the evolution of fatality counts. For the analysis we concentrated on countries with cumulative death toll of at least 1000, a number which we found to allow for a robust analysis.

### 2.3 Fatalities rescaling

In practice, not all active cases (infected individuals) are detected and reported, with the consequence that the official numbers of daily cases, and likewise the total number, is subject to under-counting. Furthermore, even when an infected individual is identified, the report is normally delayed from the moment of the infection to the occurrence of symptoms, and subsequent positive testing, a process taking up to several weeks [24,25]. Individuals identified as infected are most of the time isolated (quarantined) and the possibility that they further spread the disease is minimal. Hence, from the perspective of the outbreak dynamics, daily cases counts are an indicator for the number of individual changing from the group of infectious to the removed individuals R, which are the ones unable to spread the disease.

Miscounting is present also for official fatality counts, but to a reduced extent [26]. It is possible to estimate the extent to which the history of fatalities and infections trace each other in phase space, by comparing the functional dependence of (*X*, Δ*X*) and (*F*, Δ*F*):

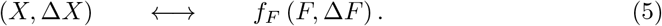

A rational for this procedure is presented within Approximate integration.

In Fig. 3 we show the relationship between daily cases and daily fatalities for the countries and US states with the highest death tolls. For some countries, like Italy, the rescaling procedure (5) works surprisingly well. The accuracy can be gauged by

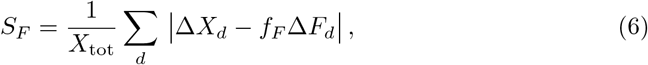

**Fig 3.**
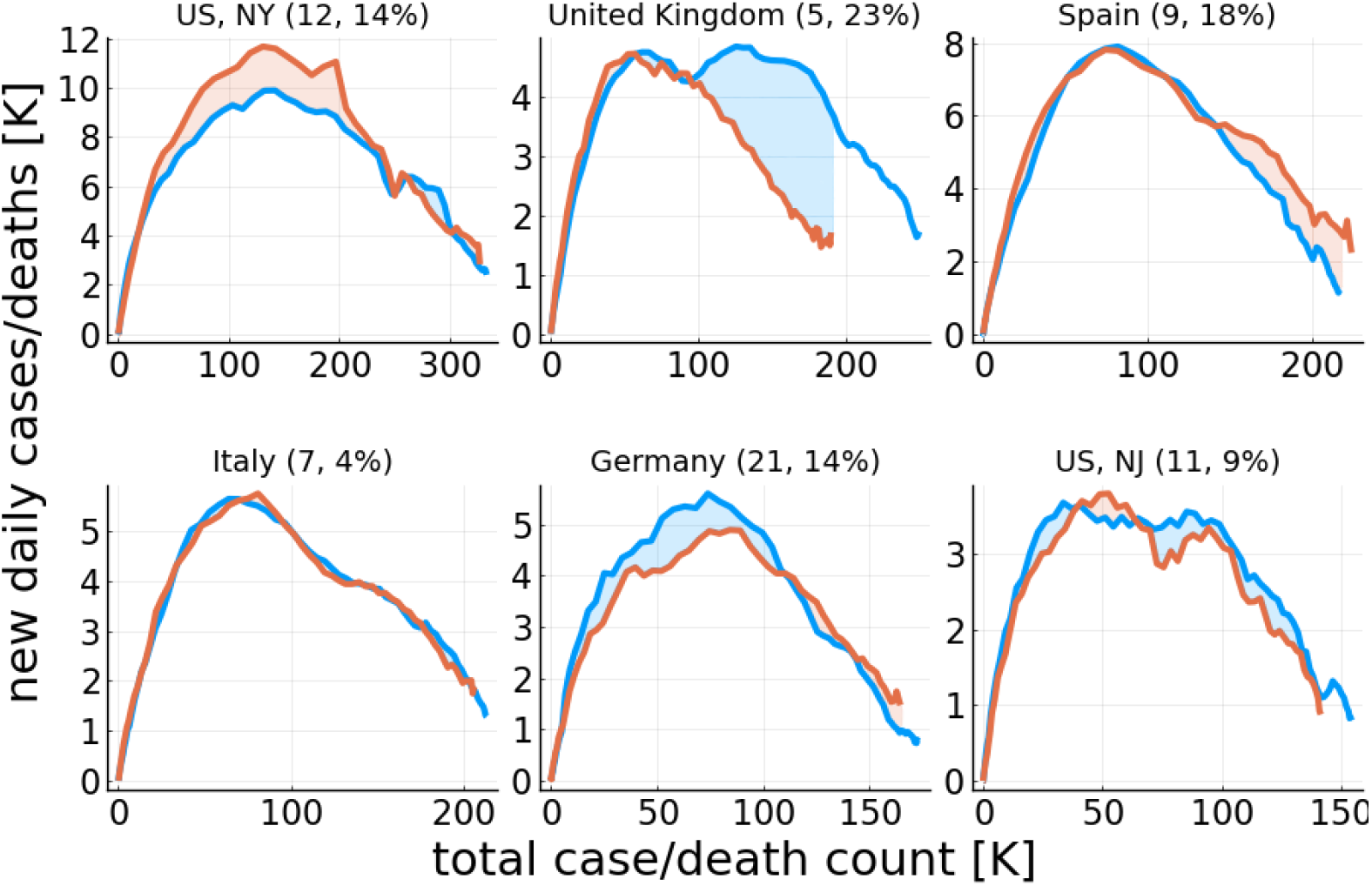
COVID-19 cases vs. fatalities. Daily new cases Δ*X* as a function of the total case count, *X* (blue) and the rescaled daily fatalities, Δ*F* → *f_F_*Δ*F*, as a function of the rescaled total death count, *F* → *f_F_F* (orange). The rescaling factors *f_F_*, given in brackets (first number), have been determined by aligning the initial slopes Δ*F*/*F* and Δ*X*/*X*. The accuracy of the rescaling, *S_F_*, measured as the relative area difference (shaded area over total area, see Eq. (6)), is given in the brackets (second number). Shown are countries and regions with the highest cumulative fatality counts, out of the ones considered here, as listed in Table 1. The data has been terminated once ΔX has fallen by 70%, which we use to define the first outbreak.

which corresponds to the percentage-wise miscounting of the daily cases Δ*X_d_* with respect to rescaled daily fatalities *f_F_*Δ*F_d_*. Note that *S_F_* → 1 when the rescaling factor *f_F_* is set to zero, since *X*_tot_ = ∑*_d_* Δ*X_d_* and |Δ*X_d_*| = Δ*X_d_*.

In Table 1 the scaling accuracies in terms of *S_F_* are listed for all countries and US state examined. Values of the order 10%-20% are typical. The quality of the matching suggest that the respective under-counting factors are stable, and not changing substantially over time. The results presented in Fig. 3 indicate also that other factors, like the success of medical treatments, seem to have changed comparatively little over the observation period, here *d* < *d′*, where the *d*′ is the cut-off date for the first peak defined in Data smoothing / peak definition.

The rescaling factors *f_F_* reported in Table 1 have been determined separately for each country and US state by aligning the initial slopes Δ*X*/*X* and Δ*F*/*F*. The rational is that the initial phase of an outbreak corresponds to the exponential-growth phase, for which the rescaling has to hold when circumstances do not change. The reason is that both case and death counts increase in the exponential phase with the same doubling time, with the delay of the fatalities contributing multiplicative to the rescaling factor *f_F_*.

**Table 1.** Model and data analyses parameters. The effective reproduction factor *g*_0_ estimated from the (*F*, Δ*F*) representation, the relative fraction *L/*(*L* + S) of long-term control, defined by Eq. (8), together with the rescaling factor *f_F_* and the accuracy *S_F_* of fatalities to case-count scaling, defined by Eq. (6). Also listed is the timescale *T_τ_* used for evaluating *F*_before_/*F*_after_ (the number of fatalities per time before/after the peak) in Fig. 5, and the cutoff date *d*′, given by the date at which the daily new cases Δ*X* of the first peak of the Covid-19 outbreak have dropped by 70% with respect to the maximum.

| Region | $g_0$ | $L/(L + S)$ | $f_F$ | $S_F$ | $T_\tau$ | $d'$ |
| --- | --- | --- | --- | --- | --- | --- |
| Canada | 1.17 | 0.35 | 11 | 0.12 | 29 | 2020-06-08 |
| France | 1.30 | 0.51 | 5 | 0.10 | 19 | 2020-04-29 |
| Spain | 1.39 | 0.32 | 9 | 0.18 | 16 | 2020-05-01 |
| Netherlands | 1.38 | 0.20 | 7 | 0.16 | 18 | 2020-05-11 |
| Germany | 1.21 | 0.49 | 21 | 0.14 | 22 | 2020-05-15 |
| Italy | 1.28 | 0.31 | 7 | 0.04 | 21 | 2020-05-06 |
| Turkey | 1.19 | 0.52 | 35 | 0.07 | 23 | 2020-05-18 |
| Portugal | 1.42 | 0.20 | 23 | 0.18 | 18 | 2020-06-05 |
| Romania | 1.13 | 0.77 | 14 | 0.12 | 29 | 2020-06-05 |
| UK | 1.29 | 0.30 | 5 | 0.23 | 20 | 2020-05-28 |
| US, NY | 1.31 | 0.43 | 12 | 0.14 | 17 | 2020-05-07 |
| US, NJ | 1.27 | 0.31 | 11 | 0.09 | 20 | 2020-05-22 |
| US, MI | 1.24 | 0.40 | 10 | 0.17 | 22 | 2020-05-22 |
| US, GA | 1.30 | 0.05 | 25 | 0.21 | 29 | 2020-06-29 |
| US, IL | 1.12 | 0.41 | 21 | 0.18 | 35 | 2020-06-24 |
| US, MA | 1.22 | 0.35 | 13 | 0.15 | 25 | 2020-06-04 |
| US, PA | 1.16 | 0.49 | 12 | 0.16 | 29 | 2020-06-06 |

### 2.4 Modeling fatality dynamics with an effective SIR model

The data presented in Fig. 3 indicates, as discussed above, that death and case counts of reported COVID-19 data rescale approximately. It is hence of interest to examine to which extent one can extract the characteristics of an outbreak directly from the fatality counts, which tend to be more reliable. For this purpose one could add a variable *F* to the SIR model and evaluate fatalities directly from first principles. Here we use the fact that *I* and *F* are necessarily related (only infected can die), modulo a time lag, which becomes however irrelevant in the XI phase-space representation. For this purpose, we use the following mapping between reported daily and total fatalities and the variables *X* = 1 − *S* and *I* of the SIR model, Eq. (1):

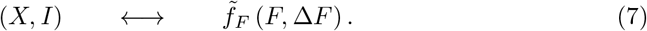

The rescaling factor defined here, 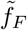, is in general different from the one used in Eq. (5). In Fig. 4 a direct comparison of the exact phase-space trajectories (*X, I*) obtained for the controlled SIR model with reported death counts is presented. To this extent the development of Δ*F* vs. *F* in phase space has been fitted using the exact solution Eq. (3) and an appropriated rescaling factor *f_F_* in Eq. (7). Note that we have estimated the free model parameters only from the data associated to the initial outbreak, for dates *d* < *d′*, as explained in Data smoothing / peak definition. The data not considered for the loss function is indicated in Fig. 4 by lighter hue markers.

**Fig 4.**
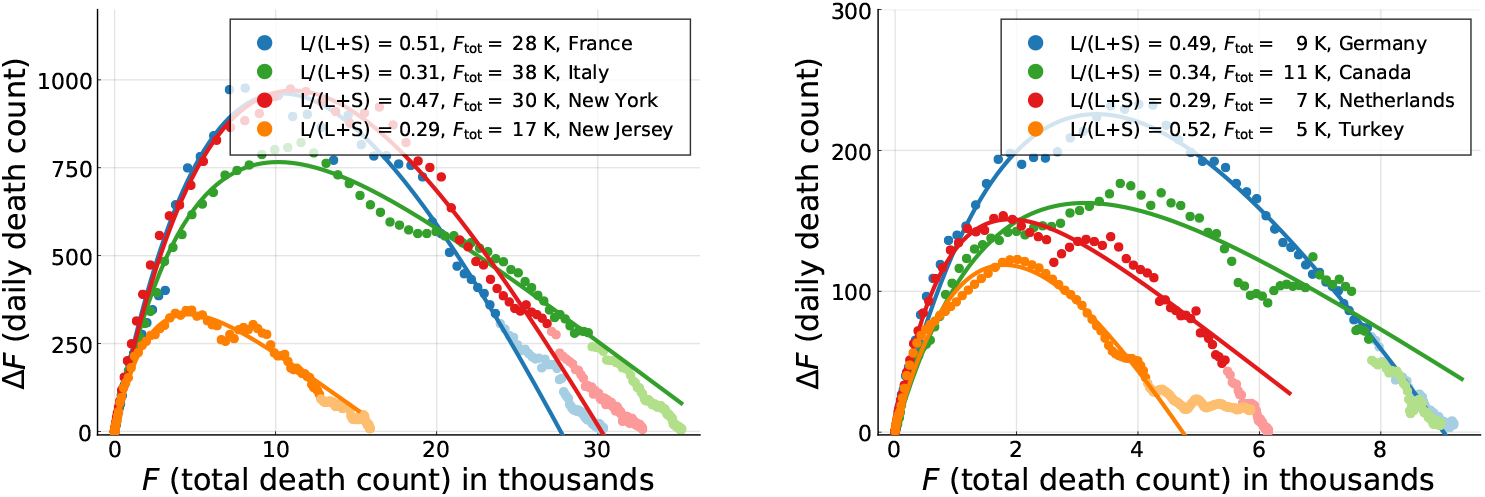
COVID-19 containment policies. Daily fatalities Δ*F* as a function of total death counts F. Comparison of data (seven-day centred averages, filled circles) and theory (lines). Points with lighter hue correspond to dates *d* > *d′* not part of the parameter estimation. The theory corresponds to optimal fits of the exact solution (16) of the controlled SIR model, Eqs. (1) and (2). The relative importance of long-term control, *L/*(*L* + S), as defined by Eq. (8), is given, together with the model based estimate of the total death toll, *F*_tot_ (assuming a single Covid-19 peak/outbreak).

Overall, the observed COVID-19 outbreaks can be described well using a mixture of long and short-term control, parametrized respectively by *α_X_* and *α_I_*. An overview of the extracted parameters is presented in Table 1. It is important to recall that the growth factor *g*_0_ of the effective SIR model used for the description of the fatality dynamics in terms of an (*X*, *I*) representation does not correspond to the medical growth factor *R*_0_. Instead, the comparison presented in Fig. 4 shows that it is possible to model the evolution of official fatality statistics directly in terms of an effective SIR model.

### 2.5 Tracing containment policies via fatality dynamics

The use of an effective SIR model to describe fatality statistics, as in Fig. 4, allows to extract containment policies, the key rational for this procedure. In absolute terms, the contributions *α_X_X* and *α_I_I* to the reduction of *g* vary strongly as function of time. We use therefore the respective values at the peak of daily fatalities, which correspond via (7) to the peak fraction *X*_peak_ of total cases, as given by (4), and to the corresponding fraction of active cases, *I*_peak_.

Hence, we use the following relation

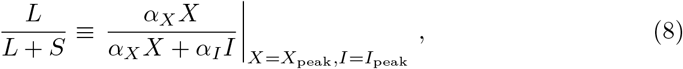

for a relative gauge, *L*/(*L* + *S*), that quantifies the fraction of control due to long-term control. Here L stays for ‘long’ and S for ‘short’. The extracted values of *L/*(*L* + *S*) are given in Table 1. For the countries shown in Fig. 4 one observes, characteristically, that the epidemics decreases fast for countries with large fractions of long-term control, and slower when short-term control dominates. Long-term control is therefore substantially more efficient in containing an epidemic outbreak.

### 2.6 Universal fatality increase after the peak

People dying of a COVID-19 infection have been typically on intensive care beforehand, which implies that the medical load is roughly proportional to the number of fatalities incurring on a daily basis. Of interest is, in this regard, whether the average medical load decreases or increase after the peak of the outbreak has been reached, in particular when averaged over a timescale *T_τ_* of several weeks.

We denote with *F*_before_, and respectively with *F*_after_, the number of deaths occurring in the *T_τ_* days before/after daily fatalities peaked, 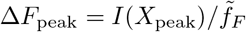, as determined by Eqs. (3) and (7). The reference period *T_τ_* is determined in our analysis by measuring the number of days that passed between *f_τ_* Δ*F*_peak_ and Δ*F*_peak_, that is between a small initial daily fatality count, *f_τ_*Δ*F*_peak_, and the peak medical load Δ*F*_peak_. See Fig. 5 for an illustration. We took *f_τ_* = 0.1 when possible, namely when the data for the same number of days after the peak was available and within the observation period. Otherwise the time span from Δ*F*_peak_ to the end of the reported timeline (or *d*′) was taken.

**Fig 5.**
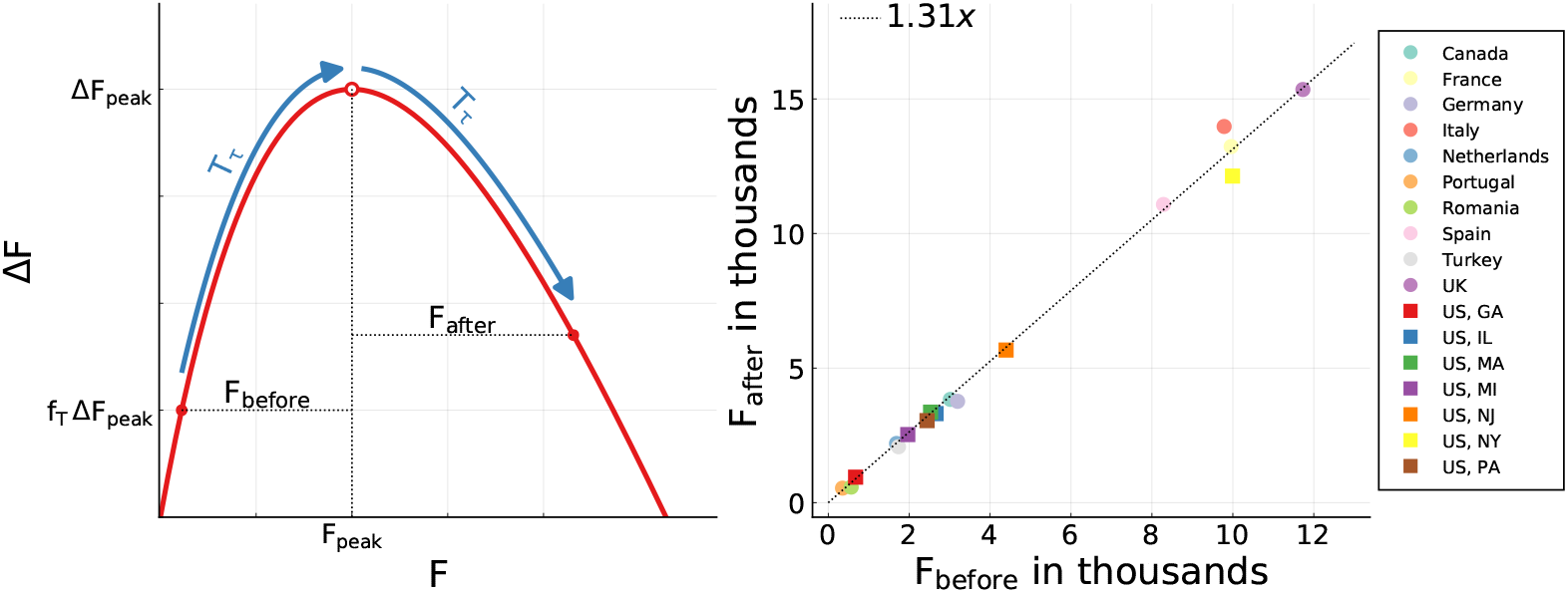
Fatalities before and after the peak – data. The observed numbers of fatalities *F*_before_ and *F*_after_ incurring over a time span *T_τ_* before/after the peak of the outbreak. *Left:* Procedure illustration. See the Methods Section for the determination of *T_τ_*. *Right:* All observed ratios *F*_after_/*F*_before_ are close to 1.3. The linear fit corresponds to a linear regression with fixed intercept (*R*^2^ = 0.994). Per time, on average 30% more death incur after the peak.

For all countries and US states examined, *F*_after_ is plotted as a function of *F*_before_ in Fig. 5. One finds a near to perfect linear relationship

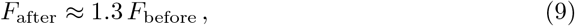

which is quite remarkable. For the linear regression with fixed intercept we find R^2^ = 0.994. It implies, that the average medical load is predictably 30% higher after the peak, than before. Given that there is a time delay between the onset of an infection and the eventual fatality, a certain increase was to be expected. The finding that this holds for a wide range of countries and regions, is however highly non-trivial. This result facilitates in our view the planning for COVID-19 specific hospital capacities. The stable relationship between medical load before and after the peak fatalities is in particular surprising in the view that the functional developments of COVID-19 outbreaks varies considerably, as illustrated in Figs. 3 and 4.

In Fig. 6 we present the ratio *X*_after_/*X*_before_ of the cumulative numbers of cases occurring in the controlled SIR model during the above defined period *T_τ_* before and after the peak. The theoretical estimates have been obtained keeping *g_0_* = 1.25 fixed

**Fig 6.**
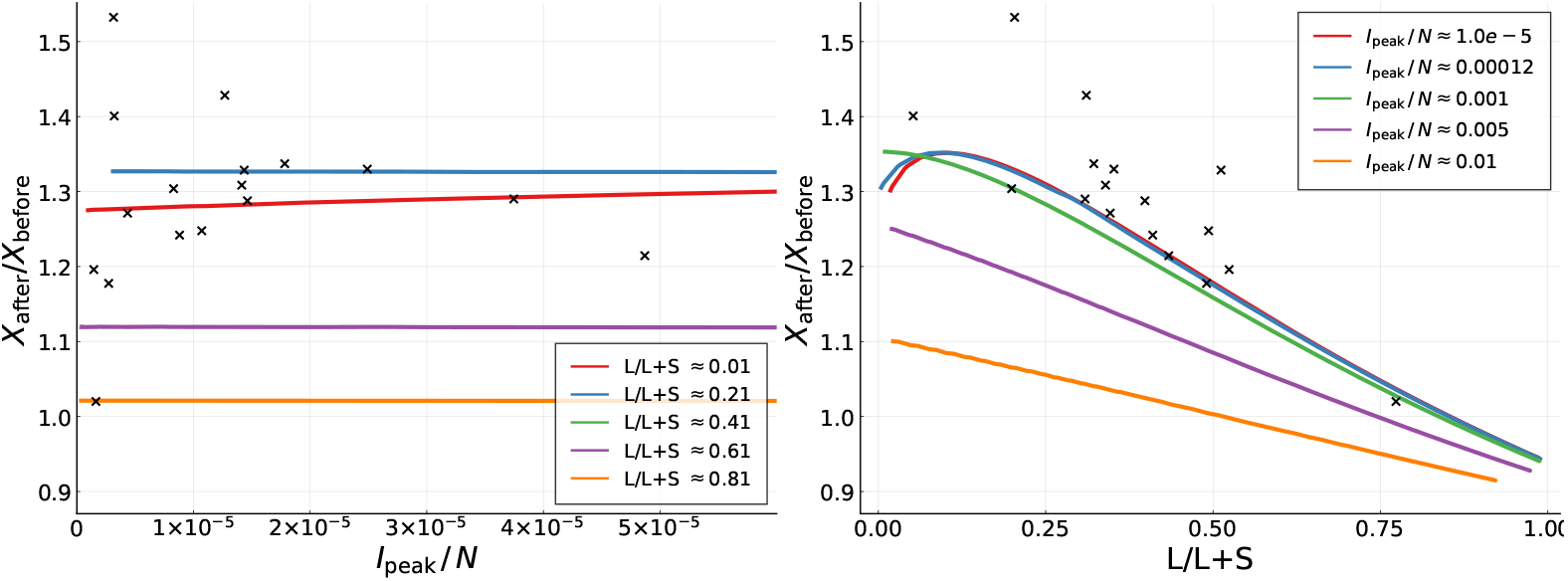
Fatalities before and after the peak – theory. For a fixed *g*_0_ = 1.25 simulation of the controlled SIR model for the ratio of average fatalities after and before the infection peak. Compare Fig. 5. A large number of simulations over a range of *α_X_* and *α_I_* have been reordered in terms of per population infection peak, *I*_peak_/*N*, and of the percentagewise contribution *L/*(*L* + *S*) of long-term control. For most countries *I*_peak_/*N* is typically of the order of 10^−4^ or smaller.

(see 1), scanning a wide range of *α_X_* and *α_I_*. Note that the field data, which is also given, scatters somewhat around the 1.3 ratio, an effect which is not as evident when using alternative representations, as in Fig. 5.

Given that case and fatality counts are related (re-scalable) for many countries, as illustrated in Fig. 3, the results presented in Fig. 6 can be understood as a first step towards an understanding why the ratio *F*_after_/*F*_before_ is of the order of 1.3 for the field data, as shown Fig. 5. In fact one observes in Fig. 6 that two conditions are necessary for *X*_after_/*X*_before_ to be of the order of 1.3, or slightly larger. Firstly, the per-population peak fraction of infected, *I_peak_*/*N* needs to be small, of the order of 10^−4^ or smaller, which is typically the case for field data. Secondly, control is dominated by short-term control, with long-term control contributing only in a minor way. This condition also holds, albeit only to a certain extent, given that *L*/(*L* + *S*) is generically smaller than 0.5; see Table 1 for details.

The data presented in Fig. 6 indicates that the size of the relative infection count and the type of containment policy enacted influence relative medical loads. Further research is however necessary to clarify why *F*_after_/*F*_before_ ≈ 1:3 holds to the observed precision.

### 2.7 Influence of initial social distancing

Google compiled changes in search-engine queries that are indicative of increasing social distancing, with an example being a reduction of inquiries concerned with traveling to the workplace. Using an average of several indicators, we compiled the Google social distancing index (GSDI); see Google social distancing index (GSDI) for details. Numerically the index is gauged with respected to its pre-Corona value. In Fig. 7 we show the correlation between the GSDI and the ratio Δ*F*/*F* between reported daily fatalities Δ*F* and total fatalities *F*. As examples we selected France, Spain, Italy, New York and New Jersey. In orders of magnitude the Google social distancing index dropped by about 80% for the European countries shown, and by about 60% for the US states. In Fig. 7f the GSDI is shown as a function of per capita fatality rates. The general trend is that the GSDI acquires somewhat lower values for European countries, with respect to US states, together with a comparative pronounced recovery. There is a certain spread in the total fatalities *F* needed for social distancing to be fully developed, as shown in Fig. 7. In per capita terms, the GSDI dropped however fast in all countries and US state examined.

**Fig 7.**
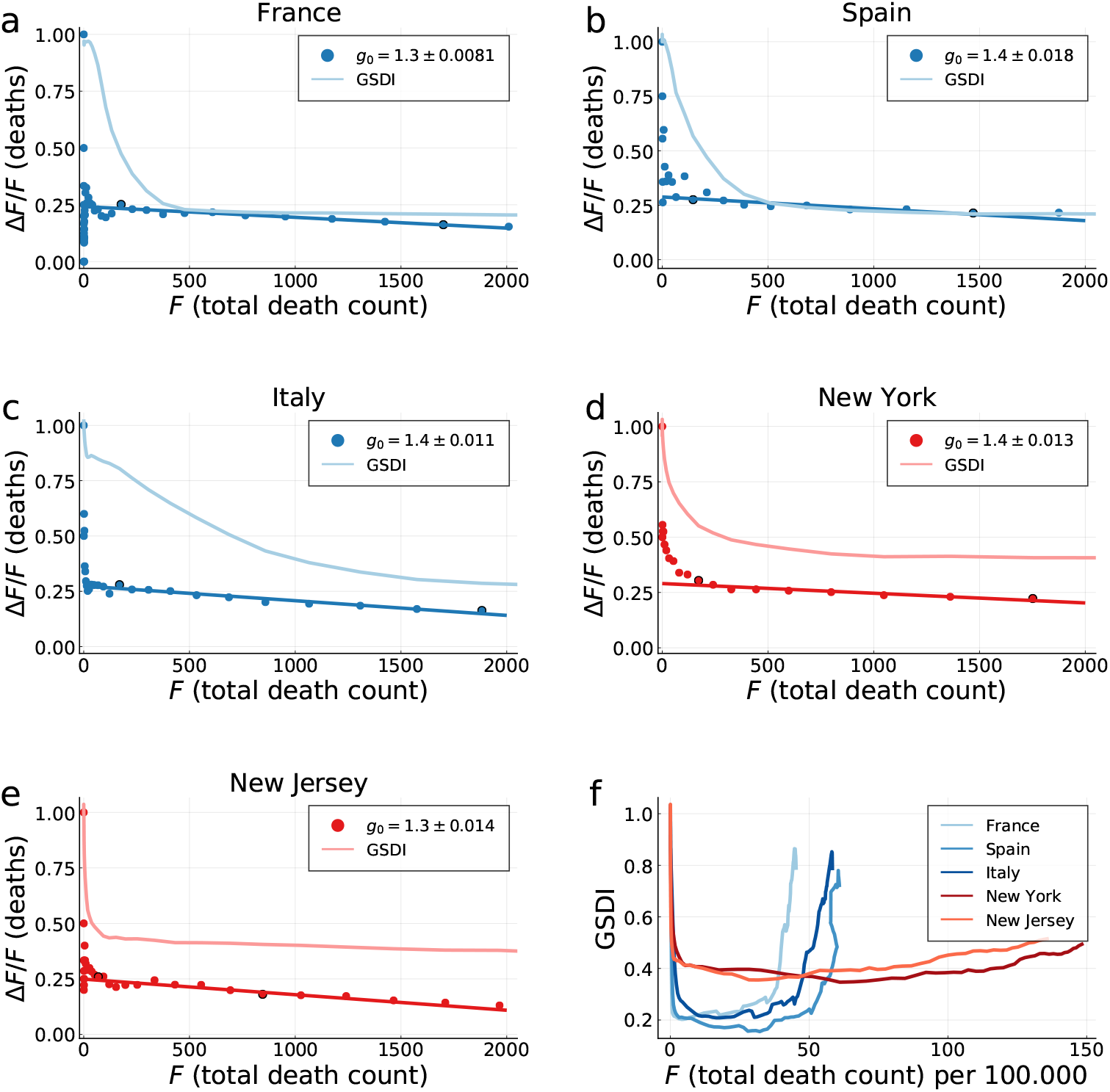
Fatalities vs. social distancing. Comparison of the Google social distancing index (GSDI), as defined in Google social distancing index (GSDI), with COVID-19 fatalities. **a)**-**e)** For France, Spain, Italy, New York and New Jersey. Shown is the ratio Δ*F*/*F* of the daily fatalities ΔF and the total death count *F* (filled circles), a linear fit between 2%-20% of the fatality peak (marked circles), and the respective GSDI. The slope has been used to calculate *g*_0_ in accordance to Eq. (21). **f)** The GSDIs on an expanded scale, now as a function of fatalities per capita (per 100,000). The selected European countries and US states show distinct behaviors.

## 3 Methods

### 3.1 Data sources

COVID-19 data sources used are the public GitHub repository of the John Hopkins Center for Systems Science and Engineering (JHU-CSSE) [27], the European Center for Disease Control open COVID-19 data (ECDC) [28], and the German Robert Koch Institute [29] (RKI). If not otherwise stated we used ECDC for country-specific data and JHU-CSSE for US states. A comparison is presented in Fig. 2.

### 3.2 Data smoothing / peak definition

The real-world epidemics reports are intrinsically noisy, with common sources of noise being report delays and under- or over-counting [30]. All data sources used show strong fluctuations with a seven day period. Accordingly, we utilize a seven-day centred moving average for data preprocessing.

Due to a multitude of errata in the data sets, it is also necessary to filter out impossible measurements, such as negative daily new cases or fatalities. As a remedy we dropped dates with negative daily fatalities Δ*F_d_* < 0. In these isolated cases the 7-day centered moving average is evaluated over the remaining seven data points, spanning eight actual days.

Endemic phases and second waves follow in most countries the initial Covid-19 outbreak, with the reason being that policies are generically relaxed when the first wave has been contained to a certain extent. The here developed framework, the controlled SIR model, is based in contrast on the assumption that containment policies, in terms of *α_X_* and *α_I_*, are constant, with the consequence that the controlled SIR model describes a single contained outbreak, and not a series of waves. However, only a well defined peak is needed for a reliable analysis, and not that the epidemic is fully eradicated. Our framework is therefore well suited to analyze the first wave of a Covid-19 epidemic, during which the containment feedback parameters *α_X_* and *α_I_* can be assumed to not change substantially.

To be specific, we define a cut-off date *d*′ as the day on which the number of daily new fatalities has fallen by 70% compared to the first peak: 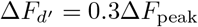. Here Δ*F*_peak_ denotes the maximum number of daily new fatalities in the 7-day centered moving average. This criterion is used to isolate the first outbreak, *d < d′*, from the subsequent course of the epidemic. The dates d^’^ used are listed in Table 1.

### 3.3 Google social distancing index (GSDI)

The Google COVID-19 mobility data describes changes in a range of mobility-related activities, each measured with respect to corresponding Google search queries [31]. We define a “Google social distancing index” (GSDI) as the the average of the three categories “workplaces”, “retail and recreation” and “transit stations”, which are given respectively by the percentage-wise activity drop relative to their pre-COVID-19 baselines. The GSDI is presented in Fig. 7.

### 3.4 Exact solution of the controlled SIR model

The phase-space trajectory of the controlled SIR model, Eqs. (1) and (2), can be derived expressively. For the derivation we extend an approach used elsewhere for the case of pure long-term control [20], starting with

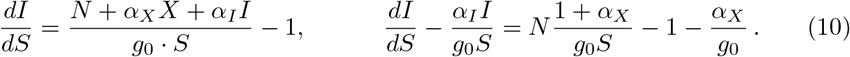

In order to obtain total differentials, one multiplies Eq. (10) with the auxiliary function

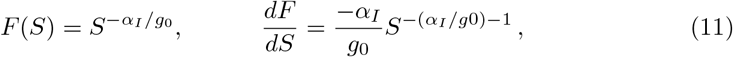

with the result

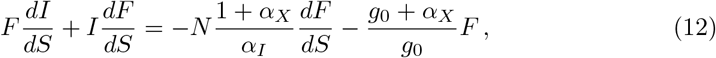

where the left-hand side is now equivalent to *d*(*FI*)*/dS*. Integration yields

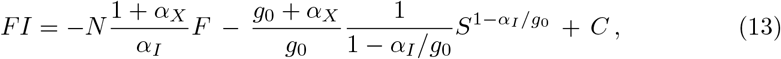

or

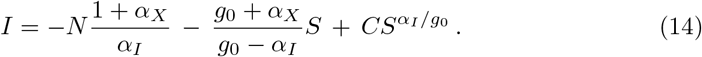

The starting condition *I*(*S* = *N*) = 0 determines the integration constant as

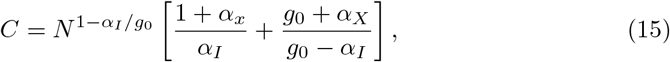

which leads to the final expression

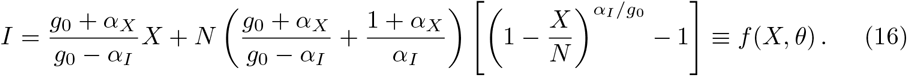

The two formal divergences on the right-hand side, *α_I_* → 0 and *α_I_* → *g*_0_, are well behaved. The first limit, *α_I_* → 0, is obtained using

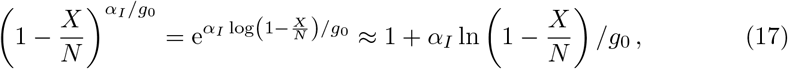

which reduces Eq. (16) to the XI representation with long-term control [20]

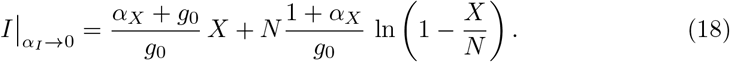

The formal divergence in the XI representation of mixed control Eq. (16) occurring when *α_I_* ≈ *g*_0_ cancels equivalently. To see this consider the expansion of

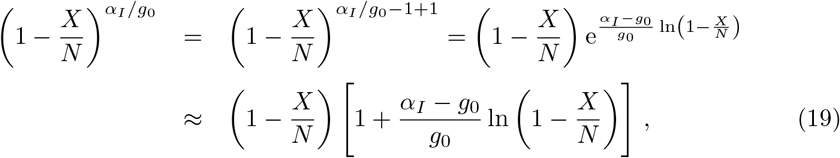

which leads to

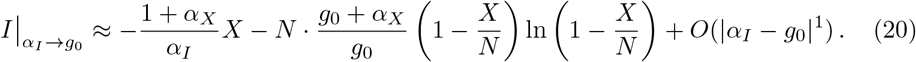

An important point is that the starting slope of Eq. (16),

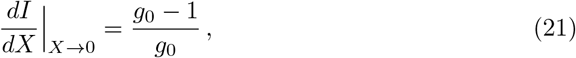

is universal, viz independent of *α_I_* and *α_X_*. For the derivation of Eq. (21) one uses 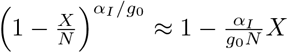. This relation has been used to calculate *g*_0_ in Fig. 7.

### 3.5 Approximate integration

Most Covid-19 datasets, contain, among other measures, the total known number of infected people *X_d_* and the total number of fatalities *F_d_* up to day *d*. For a period Δ*t* of one day, daily cases correspond to the change in total cases Δ*X_d_* = *X_d+_*_Δ_*_t_ − X_d_*.

Equivalently, daily fatalities are equal to the change in the total number of fatalities Δ*F_d_* = *F_d+_*_Δ_*_t_* − *F_d_*. We will consider the reports of daily fatalities more accurate in general than daily cases (or daily recovered) as under or over counting is less severe. In what follows we will demonstrate how one can relate daily cases and daily fatalities to the infection rate *g*(*t*), and to the XI representation.

Eq. (1) can be expressed as

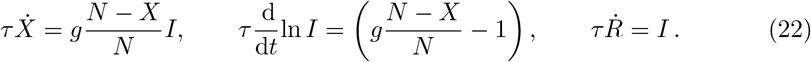

Integrating Eq. (22) between *d* and *d* + Δ*t* we obtain

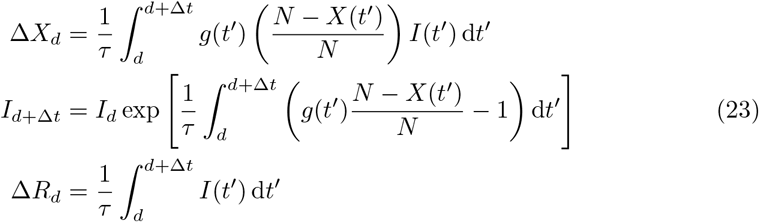

Following the approximate integration steps presented in [32], we assume that the quantities of interest, *X*(*t*), *I*(*t*), and *g*(*t*) are piecewise constant within [*d*, *d* + Δ*t*). Hence, after setting the integration interval to one day, Δ*t* = 1, we get the following set of difference equations

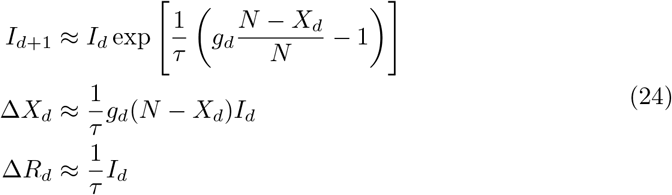

Individuals are removed, Δ*R_d_*, when recovering, or when dying, which implies Δ*F_d_* = *c_F_*Δ*R_d_*, or when quarantined, Δ*Q_d_* = *c_Q_*Δ*R_d_*. We denote here with *Q_d_* the number invididuals that are infected, but unable to infect other, either because they are quarantined at home, or because they are hospitalized. In general *c_F_* + *c_Q_* < 1 and *c_Q_* > *c_F_*. Using Eq. (24) we obtain two approximate relations for the evolution of daily quarantined and deaths,

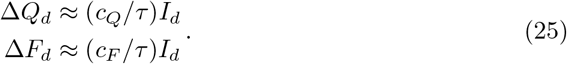

In practice, people tested positive will be advised to quarantine, or hospitalized. In view that the officially reported new cases, Δ*X_d_*, correspond to the number of positive Covid-19 tests outcomes, one has, with Eq. (25), that Δ*X_d_* ~ Δ*Q_d_* ~ *I_d_* ~ Δ*F_d_*, and hence that Δ*X_d_* scales approximately with Δ*F_d_*. We believe that this reasoning explains the observed approximate scaling between case- and death counts, as shown in Fig. 3.

As a further test of the procedure outlined above we compare in Fig. 8 the solution of the controlled SIR model with simulated data. Here we obtained *g* = *g*(*t*) by numerical integrating Eq. (1), using a 7/6 Runge-Kutta integration [33], with the phase-space representation matching the analytic expression, Eq. (3). Using Eq. (25) the timeline of infected, *I*(*t*), was used to generate simulated data for daily fatalities, Δ*F_d_*, which in turn yields the cumulative death count *F_d_ = ∑_d″_* _≤_ *_d_*Δ*F_d_*_″_. As a last step we rescaled via Eq. (7), 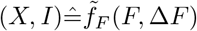, comparing the such obtained simulated data, (*X*, *I*), with the direct solution of the controlled SIR model. The agreement between the direct solution and simulated data is remarkably good.

**Fig 8.**
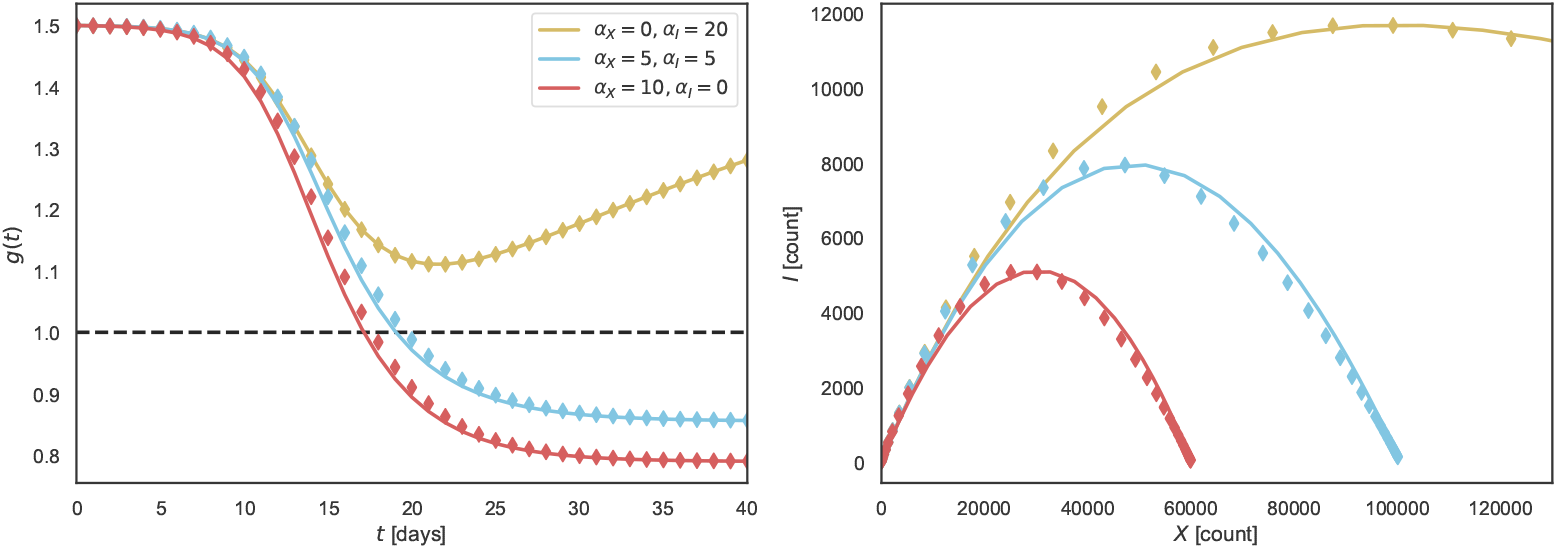
Solutions of the controlled SIR model vs. simulated data. Comparison of *g*(*t*) and *I*(*X*) obtained numerically (solid lines) and from simulated daily fatalities (diamonds). See Eq. (2) for the effective reproduction factor *g* = *g*(*t*) and Eq. (16) for the XI representation. Simulated daily and total fatalities were generated from numerically obtained active cases as Δ*F_d_* ∝ *I*(*t* = *d*), and 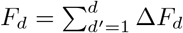. Hence diamonds correspond to a points for which 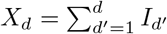. Similarly, the recovered *g_d_* are obtained from the following relation 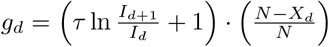. Identical initial conditions have been used for all cases, *X*(0) = 10, *I*(0) = 10, *R*(0) = 0, *g*(0) = 1.5, and *N* = 10^6^. See Approximate integration for details.

### 3.6 Parameter estimation

To fit the parameters of the controlled SIR model to the publicly available outbreak datasets, we have used the theoretical phase relation Eq. (3). The best fitting parameter values are obtained by direct minimization of the following loss function

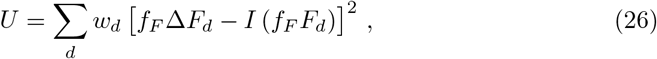

where the weights *w_d_* = *F_d_* − *F_d−_*_1_ = Δ*F_d_* ensure that long stretches of days with low fatality numbers, which are common at the beginning and the end of an epidemic, do no dominate. An optimization that weighs every data point equally, i.e. least squares, overestimates the early and in the case of one isolated outbreak the late stages of the epidemic outbreak. The rescaling factors *f_F_* used are the ones presented in Table 1.

The minimization of Eq. (26) in respect to the parameter set {*α_X_, α_I_, g*_0_} has been performed using Newton‘s method for optimization implemented in Julia by [34]. To prevent division by zero the denominators of Eq. (3) have been shifted by *εi* = 0.01*i* in the complex plane before taking the real part.

### 3.7 Simulation

The theoretical values of the ratio of average cases after and before the infection peak presented in Fig. 6 were calculated simulating the controlled SIR model Eq. (1). The numerical integration was performed using the SciMl implementation of Jim Verner’s “most efficient” 7/6 Runge-Kutta method [33]. Unless stated otherwise, the simulations were run for *τ* = 1.1 and *g*_0_ = 1.2. The population has been initialized with *I*(*t* = 0) = 10^-10^*N*. For every *α_X_* and *α_I_* we calculated the peak medical load *I*_peak_ using Eq. (4) and Eq. (3). The percentage-wise contribution *L/*(*L* + *S*) of long-term control Eq. (8) are evaluated at the point of peak infection rates.

Note that it is not possible to simulate different trajectories for which both *L*/(*L* + *S*) and *I*_peak_/*N* are fixed. For the comparison presented in Fig. 6 simulations with varying a_X_ and aj were used to bin the resulting *L*/(*L* + *S*) and *I*_peak_/*N* within about 0.5% accuracy.

## 4 Discussion

By mid 2020, the world-wide Covid-19 pandemic has entered a second phase. In most countries and regions the initial exponential growth phase has been contained to an extent that official case counts dropped substantially after the first peak. For the majority of countries and regions it is therefore possible to define an endpoint *d′* of the first wave. Here we used a simple criterion, namely a 70% drop in case numbers. Subsequent to the first wave, the development of the Corona epidemic is showing a large variety of functional dependencies.

For a large number of Covid-19 outbreaks we analyzed the first wave in terms of an effective SIR model, the controlled SIR model. The basic assumption is that containment policies can be parametrized by two parameters, *α_X_* and *α_I_*, which describe how much emphasis is placed respectively on long- and short-term control. This does not imply that containment in terms of a reduction of the basic reproduction factor is constant, but that the dependence of the reproduction factor on total and daily case counts is given by a functionally constant feedback loop. For a wide range of countries and US states we find that the official case and death counts are described well by the controlled SIR model. This observation allows to extract country-specific containment parameters, *α_X_* and *α_I_*. Containment success is found to go hand in hand with an emphasis on long-term control, with short-term control being more likely to be followed by an endemic state.

Two types of time-lines can be used to analyse Covid-19 outbreaks, one based on daily cases and another on daily fatalities. In this study we examined in particular the death toll, showing that daily and cumulative fatalities provide reliable data sources. This framework is based on the assumption that the success of medical therapies does not change substantially over the course of the observation period, here the first wave. Given the accuracy of the modelling, this assumption sees a posteriori justification, which is strengthened furthermore by the observation that case and fatality counts scale in phase-space representation, as shown in Fig. 3.

A particular interesting result of our analysis concerns the predictability of the medical load. As a measure we compare the cumulative number of fatalities over two periods of identical length, typically several weeks, just before and just after the peak of the first wave. In this regard, we find that the medical load increases on the average by 33% after the peak. This is quite a remarkable observation, in our view, given that the Covid-19 outbreaks vary substantially in between countries.

## Data Availability

All the data are available from public data repositories listed in the paper.

## Acknowledgments

We thank Daniel Gros for valuable comments and suggestions.

## Data availability

All data is available from public sources, as listed in Data sources.

